# Prevalence of Depressive Disorders and Associated Demographic Factors Among Refugees Amidst COVID-19 in Nakivale Refugee Camp in Southwestern Uganda

**DOI:** 10.1101/2020.10.21.20216754

**Authors:** Amir Kabunga, Lucas Goodgame Anyayo

## Abstract

**Introduction:** The COVID-19 pandemic and health measures to prevent it have unprecedented effects on the mental health of the refugees. However, the situation of refugees in developing countries is unclear. Thus, this study estimated the prevalence of and associated demographic factors during COVID-19 pandemic.

**The methodology:** employed a cross-sectional survey carried out in June 2020 in which 146 adult refugees from Nakivale settlement camp were randomly selected to fill out a questionnaires-demographics and PHQ9.

**Results:** revealed that the majority of respondents were female (53%) and 47% had a depressive disorder. There was no statistical relationship between demographics (gender, age and marital status) and depressive disorders.

**In Conclusion:** findings indicate that depressive disorders are prevalent among refugees in Nakivale settlement and it is important to identify refugees affected mentally and given proper treatment.

## Background

The United Nations High Commission for Refugees (UNHCR) Global Trend Report indicates that in 2018, 25.9 million refugees, 3.5 asylum seekers and 41.3 million internally displaced people have been forced to leave their homes due to armed conflicts and violence [1]. Globally, 9% of the world’s displaced people are hosted in Europe, 12% in the Americas and 84% of the world’s refugees live in developing countries [1]. Sub-Saharan Africa hosted about 26% of the world refugees [2]. Uganda, the third-largest refugee-hosting nation after Turkey (3.7 million), Pakistan (1.4 million) had 1.4 million by 2018 [1]. Many refugees besides suffering from physical injury, they too suffer mental harm [3]. However, little attention has been paid on the mental problems of the refugees.

Forced migrations are characterized by traumatic events for the refugees during migration and after settlement. They are exposed to emotionally shocking stories, images of horror and pain. Besides, they have to adapt in the host countries, different culture, unemployment and uncertainty [2]. Earlier studies show that wars and violence are associated with a greater burden of mental problems [3]–[5]. For instance studies in Nepal and the Middle East showed that the refugee respondents had mental health issues [6]. Similarly, in the Karenni refugees, anxiety and depression levels were 42% and 41% respectively [7]. Refugees may experience multiple mental issues including Post-traumatic Stress Disorder, anxiety disorder mood disorder and depression [8]. Epidemiological studies on the psychopathology of war survivors depression is one of the most frequent mental health disorders experienced by refugees [9], [10].

Demographics have often been examined as possible predictors for depression in refugee population but with inconsistent results [11]. Regarding age contradictory results have been produced, some studies reporting no association with depression [12]–[14], while others revealed a significant relationship between age and depression [15]. In a similar study in Mexico among Guatemalan refugees revealed that marital status, gender and age were related to depression [16]. Marital status and gender were related to depression among Somali refugees in Melkadida camp [17]. In another study by Bogic and colleagues [18], results showed that depression was more common among females, adults and singles. There are relatively consistent findings showing high levels of depression among females [19]-[20]. However, a study Haile and Samuel [21] found a higher prevalence rate among males than females, the results that mirrored those of Noh and colleagues [21] in Korean immigrants in Toronto.

Depression is a widespread mental problem estimated to affect about 9.8%-67.4% of refugees globally [3], [23]. Systematic reviews and meta-analysis on depression among refugees revealed variations in prevalence rates ranging between 5% and 80% [24]. One systematic review concluded that depression was one the mental health disorder among refugees with prevalence rate at 30-40% [25]. A meta-analysis reported the prevalence rate of 25-45% for depression [26]. A study in Cambodian refugees revealed that 52% of the respondents reported depression [27]. In Sub-Saharan Africa, the prevalence rate is 20% for depression in refugees [29]. A study conducted in both southern Sudan and northern Uganda found the prevalence of depression was 48% among south Sudan refugees [28]. In a similar study among urban refugees in Kampala, results showed high levels of depression disproportionately affecting women [29]. Thus the already volatile situation of refugees becomes more traumatic in the face of COVID-19 pandemic.

The COVID-19 pandemic and the health-related measures to contain it have substantially created high levels of stress all over the world [30], [31]. This unprecedented experience has multifaceted consequences for mental health and psychological wellbeing for the general population and refugees cannot be exceptional. It is now evident that COVID-19 has worsened anxiety in population leading to mental disorders including depression [31], [32]. Therefore it is important to assess peoples’ mental status in this exceptional period. Besides, early identification of the psychological disorder makes psychological interventions more effective.

The mental health status of refugees in Nakivale refugee camp, Uganda during COVID-19 is still unknown. To date, there seems to be no study assessing the prevalence of depression among Nakivale refugees during COVID-19. Besides, although studies have been conducted on demographics (age, gender and marital status) and depression, the findings are contradictory and there seems to be none among Nakivale refugees. The settlement 109,815 hosts refugees and asylum seekers from the Democratic Republic of Congo, Burundi, Somalia, Eritrea, Rwanda and Ethiopia [1]. In an attempt to address the gap we examined the burden of depression in refugees settling in in Nakivale refugee camp. We asked: What is the burden of depression among refugees during COVID-19 pandemic and are the factors associated with depression?

## Methods

### Participants

We conducted a cross-sectional survey in June 2020. We were allowed one week to collect the data required and also to avoid overcrowding within the camps. We abided by the protocols set by the Ministry of Health Uganda (MoH) task force to minimise the spread of COVID19.

The survey was carried out in Nakivale refugee settlement camp located in Southwestern Uganda and hosts refugees from Burundi, Rwanda, Somalia, Eritrea, Ethiopia and the Democratic Republic of Congo. It is one of the largest refugee settlement camps in Uganda. We conveniently sampled 146 participants from the camp that met the eligibility criteria for the study. Participants were included if they were 18 and above, those who were not sick and those who could speak at least one of the three languages-English, Kiswahili and Kinyarwanda. Participant aswere excluded if they were: below 18; were not a refugee; not staying in the camp. We obtained written consent from all study participants and followed all the guidelines and regulations set out by the Uganda National Council of Science and Technology for data collection during the pandemic as well as those of the Research Ethical Review Committee.

#### Measures

PHQ-9 is a standard tool used to screen, diagnose and monitor depressive disorders/symptoms. The tool has 9 items whose scores range from 0 (not at all) to 3 (nearly every day respectively). The 9 items represent the 9 symptoms of depression found in the DSM V[33]. In this study, we did not focus on the 10th question that identifies any subjective impairment of function. To determine the severity of the symptoms, we summed up the total of the scores of each item ticked. The categories specifying the severity of symptoms were as follows: Minimal Depression 1-4; Mild Depression 5-9; Moderate Depression 10-14; Moderately severe Depression 15-19; Severe depression 20-27. To determine the presence of a depressive disorder in a participant, we included any participant who had at least 4 symptoms that scored above 2 (except in item 9 in which any tick scoring above 1 was allowed) and either of item 1/2 or both had been ticked. We asked about demographic information including categorised age, gender and marital status of the participants. The survey questionnaire was administered by research assistants after being trained by a clinical psychologist on its proper administration. No modifications and changes were made to the questionnaire. The research team followed the standard operating procedure set out by Ministry of Health,Uganda and the Uganda National Council of Science and Technology on collection of data in the field.

#### Statistical analysis

We summarised the collected information form the questionnaire in Microsoft excel sheet which we later imported to STATA version 14.0 for statistical analysis. The predictor variables were age, gender and marital status (all categorical) while the outcome variable was depressive disorder (also categorical). We carried descriptive statistics summarizing the variables into frequency and percentages in a table. To determine the relationship, we conducted a bivariate analysis using Chi-square test of independence and Fischer’s exact test (where values were <5 in a square) with statistical significance being set at P<0.05

## Results

In this study, the prevalence of depression among refugees in Nakivale refugee settlement camp during COVID-19 was examined. Results in Table 1 indicate that over 50% (n=77) of the respondents were females and 47.3% (n=69) were males. Furthermore, majority of respondents 31.5% (n=46) were aged between 44 and 59, followed by 29.5% (n= 43) aged between 31 and 44, then 21.2% (n=31) who were aged between 18-30 and lastly 17.8% (n=26) were aged between 60 and above. The results in Table 1 show that the majority of respondents 65.8% (n=96) were married and 34.2% (n=50) were single.

**Table 1:** Demographics of the Respondents.

| Variable |  | Frequency | percentage |
| --- | --- | --- | --- |
| **Gender** | Male | 69 | 47.3 |
|  | Female | 77 | 52.7 |
| **Age** | 18-30 | 31 | 21.2 |
|  | 31-44 | 43 | 29.5 |
|  | 44-59 | 46 | 31.5 |
|  | 60 and above | 26 | 17.8 |
| **Marital status** | Married | 96 | 65.8 |
|  | Single | 50 | 34.2 |

The occurrence of depressive disorders of the respondents was recorded using PHQ-9. Table 2 reveals that 53% (n=33) of the respondents did not have depression while 47% (n=69) reported depressive disorders with females being the majority 25% (n=36).

**Table 2:** Depressive Disorders.

| Depressive disorder | Gender | Frequency | Percentage |
| --- | --- | --- | --- |
| **No Depressive Disorder** | Male | 36 | 25 |
|  | Female | 41 | 28 |
|  | **Total** | **77** | **53** |
| **With Depressive Disorder** | Male | 33 | 22 |
|  | Female | 36 | 25 |
|  | **Total** | **69** | **47** |

Further analysis was conducted to the prevalence of major depressive disorders among the respondents. Table 3 shows that of those who had a depressive disorder, 78.3% (n=54) had severe depression, with female constituting 42% (n=29 and 36.2% (n=26) being male. Also, 17.4% of the respondents with depression, had moderate depression, with again majority being female with 10.1% and 7.2% (n=5).

**Table 3:** The severity of the depressive Disorders.

| Gender | Severity of disorder |  |  |  | Total |
| --- | --- | --- | --- | --- | --- |
|  | Minimal Depression | Mild Depression | Moderate Depression | Severe Depression |  |
| Male | 0 | 3 (4.4%) | 5 (7.2%) | 25 (36.2%) | 33 (47.8) |
| Female | 0 | 0 (0.0%) | 7 (10.2%) | 29 (42.0%) | 36 (52.2%) |
|  | 0 | 3 (4.4%) | 12 (17.4) | 54 (78.3%) | 69 (100%) |

A bivariate analysis was conducted to examine the relationship between demographics and depressive disorders. Results in Table 4 indicate that there was no statistical relationship between demographics (gender, age and marital status) and outcome -depressive disorders. The gender differences had no statistical differences, the p-value (p=0.897) is greater than the alpha value (p<0.05). Age difference also revealed no statistical differences since the p-value (p=0.755) is higher than the alpha value (p<0.05). Lastly, the differences in marital status had no statistical differences because the p-value (p=0.632) is greater than the alpha value (p<0.05).

**Table 4:** Bivariate Analysis Showing relation between demographics and Depressive Disorders.

| Variable |  | P-value |
| --- | --- | --- |
| Gender | Male | 0.897 |
|  | Female |  |
| Age | 18-30 | 0.755 |
|  | 31-44 |  |
|  | 44-59 |  |
|  | 60 and above |  |
| Marital status | Married | 0.632 |
|  | Single |  |

## Discussion

COVID-19 pandemic is one of the most devastating challenging crises for public health in the modern world. Nations and different groups across the globe have suffered from a spike of agonizing psychological outcome including depression. In this context, the refugees are no exception. Thus this seems to be the first work on the prevalence of depression among refugees following COVID-19 pandemic. According to the analysis, the prevalence of depression in refugees is 47%, two thirds (25%) being women. This figure is in the range of study findings conduct in similar populations. In a study by Tamblyn et al., [35] 45% of the respondents were diagnosed with depression. Similar results (44%) were observed among Syrian asylum seekers [36]. However, it was far higher than studies conducted such as among Somali refugees in Melkadida camp (38.3%), Vietnamese refugees in America (20%) and Ethiopian immigrants (9.8%) [17], [37], [38].In this study, females reported significantly higher burden of depression compared to males. Most studies globally agree that the burden of depression is higher among women (WHO,2004). Nasıroğlu and Çeri [21] reported that females were more at risk compared to males.

The results indicate that of those who had depressive disorders, 78.3% had severe depression, with females constituting 42%. This prevalence rate is in tandem with results in refugees such as Central Africa, Middle Eastern, South-Eastern Europe and Southern Asia [41], [42]. Bandeira et al., [43] reported depression in 74% of the refugee respondents. The systematic reviews on depression among refugees results showed variations in reported prevalence rates between 5 and 80% [44]. The rate of 78.2% however, exceeded the prevalence rates other regions in studies conducted before COVID-19 such as northern Uganda, Kurdistan region of Iraq, Syria, Mexico and Lebanon ranging between 20 and 60 [10], [16], [45]–[47]. For example in the study of mahmood et al., [46] the prevalence was 59.4 for depression. The high prevalence of depression among refugees in Nakivale camp may be attributed to the effects of COVID-19 and measures to prevent or mitigate it.

The study indicates that there was no statistical relationship between depression and demographic factors (gender, age and marital status). This may not be a surprising result given that other studies on the topic have revealed contradictory results [11]. Marital status and gender were related to depression among Somali refugees in Melkadida camp [17]. In another study by Bogic et al.,[18] results showed that depression was more common among females, adults and singles. Most studies globally agree that the burden of depression is higher among women [39]). Nasıroğlu and Çeri [21] reported that females were more at risk compared to males. However, a study Haile and Samuel [37] found a higher prevalence rate among males than females, the results that mirrored those of Noh et al., [22] in Korean immigrants in Toronto.

## Limitations

The respondents’ recruitment was restricted to only refugees who were in the camp, speaking at least one of the 3 languages-English, Kiswahili and Kinyarwanda. We may not generalise the findings to all refugees. Besides this was a cross-sectional study with its limitations thus a causal effect relationship is not possible.

## Conclusion

COVID-19 pandemic has caused several mental health problems notably Depression. The results in this study indicate high levels of depressive disorders among the refugees. Therefore amidst the pandemic, it is important to identify refugees affected mentally and given proper treatment. This study adds to the existing literature on depression by showing evidence of elevated rates of depression during COVID-19. The overall high prevalence of depression in Nakivale refugee settlement camp proves the need for the urgency of screening and treatment of depression among the refugees. According to WHO, depression among refugees needs immediate attention and policies to deal with it [1].

## Data Availability

data for the research is available on request from the corresponding author.
can be contacted by email on.
When it has been deposited in the repository, a notification will be sent to medRxiv

## Acknowledgement

The authors extend their heartfelt thanks to the MoH for their support during data collection amidst the COVID-19 pandemic. We would also wish to thank students of Lira University for their contribution to data collection and management.

## Authors contribution

AK conceived the study. Both authors designed and conducted the study. LA contributed to data analysis. Both authors contributed to results and review of the manuscript. They too approved the final version and publication of the manuscript.

## Availability of data and materials

Data set to support the conclusions are available and a data file is attached with this submission.

## Ethical approval and consent

The study was approved by the National HIV/AIDS Research Committee of Uganda National Council of Science and Technology - FAN: IREC0001698. Access to the camp was granted by the office of the Prime Minister and the MoH-Uganda through the district COVID-19 task force. We obtained written informed consent from every participant that took part in the study. A clinical Psychologist and a Psychiatrist were on standby in cases where a participant needed psychosocial and psychiatric support or services.

## References

[1] UNHCR, ‘Uganda Country Refugee Response Plan’, UN Refug. Agency, no. Jan 2019-Dec 2020, p. 72, 2019, [Online]. Available: http://reporting.unhcr.org/sites/default/files/UgandaCountryRRP2019-20%28January2019%29.pdf.

[2] K. Triantafyllou et al., ‘Mental health and psychosocial factors in young refugees, immigrants and Greeks: A retrospective study.’, Psychiatr. Psychiatr., vol. 29, no. 3, p. 231, 2018.

[3] D. Teodorescu, T. Heir, E. Hauff, T. Wentzel□Larsen, and L. Lien, ‘Mental health problems and post□migration stress among multi□traumatized refugees attending outpatient clinics upon resettlement to Norway’, Scand. J. Psychol., vol. 53, no. 4, pp. 316–332, 2012.

[4] E. Goldmann and S. Galea, ‘Mental health consequences of disasters’, Annu. Rev. Public Health, vol. 35, pp. 169–183, 2014.

[5] J. Ben Farhat et al., ‘Syrian refugees in Greece: experience with violence, mental health status, and access to information during the journey and while in Greece’, BMC Med., vol. 16, no. 1, p. 40, 2018.

[6] M. J. D. Jordans, M. Semrau, G. Thornicroft, and M. van Ommeren, ‘Role of current perceived needs in explaining the association between past trauma exposure and distress in humanitarian settings in Jordan and Nepal’, Br. J. Psychiatry, vol. 201, no. 4, pp. 276–281, 2012.

[7] L. A. Vonnahme, E. W. Lankau, T. Ao, S. Shetty, and B. L. Cardozo, ‘Factors associated with symptoms of depression among Bhutanese refugees in the United States’, J. Immigr. Minor. Heal., vol. 17, no. 6, pp. 1705–1714, 2015.

[8] J. T. V. M. De Jong, I. H. Komproe, and M. Van Ommeren, ‘Common mental disorders in postconflict settings’, Lancet, vol. 361, no. 9375, pp. 2128–2130, 2003.

[9] N. Morina, K. Stam, T. V Pollet, and S. Priebe, ‘Prevalence of depression and posttraumatic stress disorder in adult civilian survivors of war who stay in war-afflicted regions. A systematic review and meta-analysis of epidemiological studies’, J. Affect. Disord., vol. 239, pp. 328–338, 2018.

[10] C. Acarturk, M. Cetinkaya, I. Senay, B. Gulen, T. Aker, and D. Hinton, ‘Prevalence and predictors of posttraumatic stress and depression symptoms among Syrian refugees in a refugee camp’, J. Nerv. Ment. Dis., vol. 206, no. 1, pp. 40–45, 2018.

[11] Y. Nesterko, D. Jäckle, M. Friedrich, L. Holzapfel, and H. Glaesmer, ‘Factors predicting symptoms of somatization, depression, anxiety, post-traumatic stress disorder, self-rated mental and physical health among recently arrived refugees in Germany’, Confl. Health, vol. 14, no. 1, pp. 1–12, 2020.

[12] M. C. Chung et al., ‘The impact of trauma exposure characteristics on post-traumatic stress disorder and psychiatric co-morbidity among Syrian refugees’, Psychiatry Res., vol. 259, pp. 310–315, 2018.

[13] E. Georgiadou, A. Zbidat, G. M. Schmitt, and Y. Erim, ‘Prevalence of mental distress among Syrian refugees with residence permission in Germany: a registry-based study’, Front. psychiatry, vol. 9, p. 393, 2018.

[14] S. J. Song, A. Subica, C. Kaplan, W. Tol, and J. de Jong, ‘Predicting the mental health and functioning of torture survivors’, J. Nerv. Ment. Dis., vol. 206, no. 1, pp. 33–39, 2018.

[15] R. V Reed, M. Fazel, L. Jones, C. Panter-Brick, and A. Stein, ‘Mental health of displaced and refugee children resettled in low-income and middle-income countries: risk and protective factors’, Lancet, vol. 379, no. 9812, pp. 250–265, 2012.

[16] M. Sabin, B. L. Cardozo, L. Nackerud, R. Kaiser, and L. Varese, ‘Factors associated with poor mental health among Guatemalan refugees living in Mexico 20 years after civil conflict’, Jama, vol. 290, no. 5, pp. 635–642, 2003.

[17] F. Feyera, G. Mihretie, A. Bedaso, D. Gedle, and G. Kumera, ‘Prevalence of depression and associated factors among Somali refugee at melkadida camp, southeast Ethiopia: a crosssectional study’, BMC Psychiatry, vol. 15, no. 1, p. 171, 2015.

[18] M. Bogic, A. Njoku, and S. Priebe, ‘Long-term mental health of war-refugees: a systematic literature review’, BMC Int. Health Hum. Rights, vol. 15, no. 1, p. 29, 2015.

[19] S. Nasıroğlu and V. Çeri, ‘Posttraumatic stress and depression in Yazidi refugees’, Neuropsychiatr. Dis. Treat., vol. 12, pp. 2941–2948, 2016, doi: 10.2147/NDT.S119506.

[20] de J. J. Song SJ, Subica A, Kaplan C, Tol W, ‘Predicting the Mental Health and Functioning of Torture Survivors’, J Nerv Ment Dis, vol. 206, no. 1, pp. 33–39, 2018.

[21] S. N. Haile Fenta, Ilene Hyman, ‘Determinants of depression among Ethiopian immigrants and refugees in Toronto’, J. Nerv. Ment. Dis., vol. Volume 192, no. 5, pp. 363–372, 2004.

[22] S. Noh, Z. Wu, M. Speechley, and V. Kaspar, ‘Depression in Korean immigrants in Canada: II. Correlates of gender, work, and marriage.’, J. Nerv. Ment. Dis., 1992.

[23] G. N. Marshall, T. L. Schell, M. N. Elliott, S. M. Berthold, and C.-A. Chun, ‘Mental health of Cambodian refugees 2 decades after resettlement in the United States’, Jama, vol. 294, no. 5, pp. 571–579, 2005.

[24] J.-R. Henkelmann et al., ‘Anxiety, depression and post-traumatic stress disorder in refugees resettling in high-income countries: systematic review and meta-analysis’, BJPsych Open, vol. 6, no. 4, 2020.

[25] G. Turrini, M. Purgato, F. Ballette, M. Nosè, G. Ostuzzi, and C. Barbui, ‘Common mental disorders in asylum seekers and refugees: umbrella review of prevalence and intervention studies’, Int. J. Ment. Health Syst., vol. 11, no. 1, p. 51, 2017.

[26] J. Lindert, O. S. von Ehrenstein, S. Priebe, A. Mielck, and E. Brähler, ‘Depression and anxiety in labor migrants and refugees–a systematic review and meta-analysis’, Soc. Sci. Med., vol. 69, no. 2, pp. 246–257, 2009.

[27] S. M. Lingala and M. G. M. Mhs. Ghany, ‘毛细血管 网 来源的米色脂肪和它节老鼠的代谢内稳态 HHS Public Access’, vol. 25, no. 3, pp. 289–313, 2016, doi:110.1016/j.bbi.2017.04.008.

[28] U. K. Karunakara et al., ‘Traumatic events and symptoms of post-traumatic stress disorder amongst Sudanese nationals, refugees and Ugandans in the West Nile’, Afr. Health Sci., vol. 4, no. 2, pp. 83–93, 2004.

[29] C. H. Logie, M. Okumu, S. Mwima, R. Hakiza, D. Chemutai, and P. Kyambadde, ‘Contextual factors associated with depression among urban refugee and displaced youth in Kampala, Uganda: findings from a cross-sectional study’, Confl. Health, vol. 14, no. 1, pp. 1–13, 2020.

[30] J. Lai et al., ‘Factors associated with mental health outcomes among health care workers exposed to coronavirus disease 2019’, JAMA Netw. open, vol. 3, no. 3, pp. e203976–e203976, 2020.

[31] W. Cao et al., ‘The psychological impact of the COVID-19 epidemic on college students in China’, Psychiatry Res., p. 112934, 2020.

[32] Y.-T. Xiang et al., ‘Timely mental health care for the 2019 novel coronavirus outbreak is urgently needed’, The Lancet Psychiatry, vol. 7, no. 3, pp. 228–229, 2020.

[33] A. P. Association, Diagnostic and statistical manual of mental disorders (DSM-5®). American Psychiatric Pub, 2013.

[34] J. M. Tamblyn, A. J. Calderon, S. Combs, and M. M. O’Brien, ‘Patients from abroad becoming patients in everyday practice: torture survivors in primary care’, J. Immigr. Minor. Heal., vol. 13, no. 4, pp. 798–801, 2011.

[35] O. M. Tamblyn JM, Calderon AJ, Combs S, ‘Patients from abroad becoming patients in everyday practice: torture survivors in primary care’, J Immigr Minor Heal., vol. 13, no. 4, pp. 798–801, 2011, doi:10.1007/s10903-010-9429-2. PMID: pudmed: 21188531.

[36] T. B. Danielle N. Poole, Bethany Hedt-Gauthier, Shirley Liao, Nathaniel A. Raymond, ‘Major depressive disorder prevalence and risk factors among Syrian asylum seekers in Greece’, BMC Public Health, vol. 18, no. 908, pp. 2–9, 2018, doi: https://doi.org/10.1186/s12889-018-5822-x.

[37] H. Fenta, I. Hyman, and S. Noh, ‘Determinants of depression among Ethiopian immigrants and refugees in Toronto’, J. Nerv. Ment. Dis., vol. 192, no. 5, pp. 363–372, 2004.

[38] D. N. Poole, B. Hedt-Gauthier, S. Liao, N. A. Raymond, and T. Bärnighausen, ‘Major depressive disorder prevalence and risk factors among Syrian asylum seekers in Greece’, BMC Public Health, vol. 18, no. 1, p. 908, 2018.

[39] W. H. Organization, W. H. O. S. A. Department, W. H. O. D. of M. Health, and S. Abuse, Global status report on alcohol 2004. World Health Organization, 2004.

[40] S. Nasıroğlu and V. Çeri, ‘Posttraumatic stress and depression in Yazidi refugees’, Neuropsychiatr. Dis. Treat., vol. 12, p. 2941, 2016.

[41] S. J. Song, C. Kaplan, W. A. Tol, A. Subica, and J. de Jong, ‘Psychological distress in torture survivors: pre-and post-migration risk factors in a US sample’, Soc. Psychiatry Psychiatr. Epidemiol., vol. 50, no. 4, pp. 549–560, 2015.

[42] C. C. Schubert and R.-L. Punamäki, ‘Mental health among torture survivors: cultural background, refugee status and gender’, Nord. J. Psychiatry, vol. 65, no. 3, pp. 175–182, 2011.

[43] M. Bandeira, C. Higson-Smith, M. Bantjes, and P. Polatin, ‘The land of milk and honey: a picture of refugee torture survivors presenting for treatment in a South African trauma centre.’, Torture Q. J. Rehabil. torture Vict. Prev. torture, vol. 20, no. 2, pp. 92–103, 2010.

[44] L. H. U. Bustamante, R. O. Cerqueira, E. Leclerc, and E. Brietzke, ‘Stress, trauma, and posttraumatic stress disorder in migrants: a comprehensive review’, Brazilian J. Psychiatry, vol. 40, no. 2, pp. 220–225, 2018.

[45] F. Kazour et al., ‘Post-traumatic stress disorder in a sample of Syrian refugees in Lebanon’, Compr. Psychiatry, vol. 72, pp. 41–47, 2017.

[46] H. N. Mahmood, H. Ibrahim, K. Goessmann, A. A. Ismail, and F. Neuner, ‘Post-traumatic stress disorder and depression among Syrian refugees residing in the Kurdistan region of Iraq’, Confl. Health, vol. 13, no. 1, pp. 1–11, 2019.

[47] J. Mugisha, H. Muyinda, S. Malamba, and E. Kinyanda, ‘Major depressive disorder seven years after the conflict in northern Uganda: burden, risk factors and impact on outcomes (The Wayo-Nero Study)’, BMC Psychiatry, vol. 15, no. 1, p. 48, 2015.

